# Significance and clinical suggestions for the Somatosensory Evoked Potentials increased in amplitude revealed by a large sample of neurological patients

**DOI:** 10.1101/2022.04.12.22273791

**Authors:** Davide Rossi Sebastiano, Daniele Cazzato, Elisa Visani, Eleonora Dalla Bella, Laura Brambilla, Grazia Devigili, Paola Caroppo, Lorenzo Maggi, Lorenzo Nanetti, Ettore Salsano, Laura Canafoglia, Isabella Canavero, Elena Anghileri, Deborah Bonfoco, Paola Lanteri

## Abstract

**Objectives:** To investigate the relationship between N20-P25 peak-to-peak amplitude (N20p-P25p) of Somatosensory Evoked Potentials (SEPs) and the occurrence of abnormalities of the peripheral and/or central sensory pathways and of myoclonus/epilepsy, in 308 patients with increased SEPs amplitude from upper limbs stimulation

**Methods:** We compared cortical response (N20p-P25p) in different groups of patients identified by demographic, clinical and neurophysiological factors and performed a cluster analysis for classifying the natural occurrence of subgroups of patients.

**Results:** No significant differences of N20p-P25p were found among different age-dependent groups, and in patients with or without PNS/CNS abnormalities of sensory pathways, while myoclonic/epileptic patients showed higher N20p-P25p than other groups. Cluster analysis identified four clusters including patients with myoclonus/epilepsy, patients with central sensory abnormalities, patients with peripheral sensory abnormalities, patients with neither myoclonus nor sensory abnormalities.

**Conclusions:** Increased N20p-P25p correlated to different pathophysiological conditions: strong cortical hyperexcitability in patients with cortical myoclonus and/or epilepsy and enlarged N20p-P25p, while milder increase of N20p-P25p could be underpinned by plastic cortical changes following abnormalities of sensory pathways, or degenerative process involving the cortex. SEPs increased in amplitude cannot be considered a specific correlated for myoclonus/epilepsy, but it in several neurological disorders may represent a sign of adaptive, plastic and/or degenerative cortical changes.

## INTRODUCTION

The Somatosensory Evoked Potentials (SEPs) are a well-established neurophysiological technique for the evaluation of the somatosensory pathways from peripheral nerve to cortex.[1] Without considering their role in the intensive care unit,[2] SEPs are largely used in the diagnostic workup of the neurological diseases affecting the Central Nervous System (CNS)[1] and, albeit some cautions, even in those affecting the Peripheral Nervous System (PNS), as a complementary tool of ElectroNeuroGraphy (ENG), in revealing the abnormalities of the proximal tract of PNS.[3]

First described by Dowson,[4] SEPs increased in amplitude (SEPIA), the so-called “giant SEPs”, have been usually related to cortical myoclonus and especially to Progressive Myoclonic Epilepsies (PMEs).[5-7] Among others, giant SEPs were reported in anoxic brain injury,[8] epilepsy,[9] Creutzfeldt-Jakob Disease (CJD)[10] and neuronal ceroid lipofuscinosis [11]. Our group contributed to clarify the relationship between cortical hyperexcitability, reflex myoclonus and SEPIA.[12-13]

However, the finding of SEPIA without any evidence of myoclonus or epilepsy is not uncommon in the daily clinical practice, albeit “sporadic” in literature, as suggested by few studies on Multiple System Atrophy (MSA),[14] Progressive Supranuclear Palsy (PSP),[15] Autosomal Recessive Cerebellar Ataxia type 3,[16] Amyotrophic Lateral Sclerosis (ALS) and Motor Neuron Diseases (MND),[17]. Especially two recent works [18,19] reopened the debate on the SEPIA and their clinical significance: the vast majority [19] or all of the patients [18] included in these samples suffered from various non-epileptic and non-myoclonic disorders, implicitly suggesting that SEPIA are not exclusively related neither to epilepsy and/or reflex cortical myoclonus not to a definite disease. Unfortunately, it is still poorly understood the relationship between SEPIA and the abnormalities of central and peripheral sensory pathways and the presence of myoclonus and epilepsy, as well as the possible significance in the daily clinical practice of SEPIA in patients without myoclonus or epilepsy.

Hence, we retrospectively evaluated all the SEPs performed in an extended sample of adult patients collected in our laboratory in the latest 2.5 years. Data of SEPs were related to diagnosis, presence/absence of sensory conduction abnormalities and presence/absence of myoclonus/epilepsy. Finally, to explore natural subgroups in our cohort, we performed a cluster analysis to establish particular grouping of individuals based on their features.

## METHODS

### Patients and data collection

We retrospectively evaluated all (2365) the SEPs performed for diagnostic purposes in our laboratory at the Neurological Institute “Carlo Besta” of Milan, from 2019, January 1^st^, to 2021, June 30^th^.

We included all the people with age > 14 years who completed SEPs examination in standard laboratory setting, with the exception of patients with disorder of consciousness and/or in intensive care unit. We identified patients with earlier cortical response for upper limbs stimulation (N20p-P25p) exceeding 7.44 µV in at least one side of stimulation, i.e., the mean value + 3 standard deviations (SD) of our internal normative data for SEPs evoked from contralateral median nerve stimulation (normative value of N20p-P25p 4.26±1.06 µV) [1,25]. Patients were included in the sample only once.

For all the patients selected we collected the following data: age, diagnosis and diagnostic group (see below), presence/absence of cortical myoclonus and/or epilepsy and of abnormalities of CNS/PNS sensory conduction, N20p-P25p for both side of stimulation.

#### Diagnosis and diagnostic groups

All the patients were organized in 16 groups including different neurological diseases (table 1), i.e. autoimmune disorders affecting CNS (CNS-IMM); leucoencephalopaties (CNS-LEUCO); CNS tumors (CNS-TUM); vascular encephalopathies (CNS-VASC); cognitive disorders (COG.DIS); epileptic and/or myoclonic syndromes (EPI-MYO); hereditary spastic paraparesis (HSP); motor neuron diseases (MND); myopathies and disorders of the neuromuscular junction (MYOP); neuropathies (NEUROP); Parkinson disease (PD), parkinsonisms and related disorders (PARK), spino-cerebellar ataxia (SCA); disorders affecting the spine (SPINE). Patients affected by a neurological disease not included in the previous lists were included in “other neurological disease” (OTHER-NEU); patients with functional symptoms without definite neurological diseases/syndromes were included in the group of “non-organic” (NON-ORGANIC); patients without a definite diagnosis or with two or more neurological diseases/syndromes were included in “unknown/undetermined” (UNK/UND).

**Table 1.** Overview of the diagnosis for all patients and their inclusion into groups.

| **Etiological group (N patients)** |  |
| --- | --- |
| CNS-IMM (30) | Autoimmune Encephalitis (2), Autoimmune Limbic Encephalitis (1), Devic Syndrome (2), RR-Multiple Sclerosis (16), PP-Multiple Sclerosis (1), SP- Multiple Sclerosis (1), Myelitis (5), Isolated Optic Neuritis (2) |
| CNS-LEUCO (12) | Adrenomyeloneuropathy (2), Alexander disease (1), Adrenoleucodystrophy (7), Undetermined leucoencephalopathy (2) |
| CNS-TUM (13) | Ependymoma (3), Meningioma (3), Metastasis (3), Ependimoma (3), Neurinoma (1) |
| CNS-VASC (7) | Vascular encephalopathy [ischemic] (3), Lacunar stroke [ischemic] (2), Hemorrhagic stroke (2) |
| COG DIS (23) | Alzheimer disease and mixed dementia (3), Fahr Syndrome (1), Fronto-temporal dementia(5), Idiopathic normal-pressure hydrocephalus (3), Lewy-Body Dementia (1), Prion disease-CJD (8), Prion disease-FFI (1), Prion disease-GSS (1) |
| EPI-MYO (20) | 15q11.2 duplication (1), Aicardi syndrome (1), Neuronal ceroid lipofuscinosis-3 (1), Progressive myoclonic epilepsy type 1 (2), Jeavons syndrome (1), Juvenile myoclonic epilepsy (1), KCNA1 mutation (1), KCNA2 mutation (1), SCNA1 mutation (1), Sialidosis (1), Undetermined epileptic/myoclonic syndrome (9) |
| HSP (10) | KIF1A-mutation (1), Spastic Paraplegia type-4 (2), Spastic Paraplegia type-5 (1), Undetermined hereditary progressive paraparesis (6) |
| MND (50) | Genetic forms of MND – ARGHX (1), Genetic forms of MND – C9ORF72 (2), Genetic forms of MND – OPTN (1), Genetic forms of MND – RUNX2 (1), Genetic forms of MND – SOD1 (2), Genetic forms of MND – TARDBP (1), Genetic forms of MND – UBQLN2 (1), Sporadic- Bulbar onset ALS (8), Sporadic- Limb onset ALS (24), Post-polio Syndrome (1), Primary Lateral Sclerosis (7) |
| MYOP (13) | Myotonic dystrophy type 1 (1), Inclusion body myositis (1), Myasthenia gravis (3), |
|  | Stiff-person syndrome (3), Undetermined myopathy (5) |
| NEUROP (18) | Brachial plexus neuritis (2), Alcoholic neuropathy (1), Anti-MAG peripheral neuropathy (1), Diabetic neuropathy (2), Hereditary sensory-motor neuropathy (2), MGUS related neuropathy (1), Amyloidosis (1), Vincristine-induced peripheral neuropathy (1), Small fiber neuropathy (1), Postherpetic neuralgia (1), Undetermined neuropathy (5) |
| PARK (28) | Corticobasal degeneration (4), Multiple system atrophy (14), Parkinson Disease (3), Progressive Supranuclear Palsy (4), Vascular Parkinsonism (2), Undetermined parkinsonism (1) |
| SCA (8) | Autosomal recessive cerebellar ataxia 2 (1), CANVAS (1), Undetermined spino-cerebellar ataxia (6) |
| SPINE (32) | Chiari Malformation type 1 (2), Spinal Arteriovenous Fistula (2), Discal herniation [cervical] (1), Discal herniation [lumbar] (2), Hirayama disease (3), Traumatic injury (2), Spinal stenosis (20) |
| OTHER-NEU (20) | AMPPE (1), Bechet disease (1), Dystonia (2), Fibromyalgia (2), Headache (4), Neurosyphilis (1), Combined sclerosis [carential] (3), Paraneoplastic neurological syndromes (3), Essential tremor (3) |
| NON-ORGANIC (9) | Non-organic disorders |
| UNK/UND (15) | Unknown or undetermined etiology |
| Legend: RR = Relapsing–Remitting; PP = primary progressive; SP = secondary progressive; CJD = Creutzfeldt-Jakob disease; FFI = Fatal familial insomnia; GSS= Gerstmann-Sträussler-Scheinker disease; MND = Motor Neuron Disease; ALS = Amyotrophic Lateral Sclerosis; MGUS = Monoclonal Gammopathy of Undetermined Significance; CANVAS = Cerebellar Ataxia with Neuropathy and bilateral Vestibular Areflexia Syndrome; AMPPE = Acute multifocal placoid pigment epitheliopathy |  |

#### Somatosensory Evoked Potentials

SEPs examination were performed according to the recommendations for the clinical use of SEPs.[1] Data were collected with Nicolet EDX system, Natus Neurology, Middleton, WI, USA, at sampling frequency of 2 kHz and filtered in 5–2000 Hz band. The value of the N20p-P25p for the upper limbs SEPs was collected for the subsequent analysis.

Two clinical neurophysiologists (DRS and DC) revised SEPs and, in 201 of the total 308 patients, ENG, in order to assess abnormalities of the CNS and/or PNS sensory pathway. The raters independently evaluated SEPs and ENG, classifying them in a dichotomic way, i.e. presence or absence of CNS and/or PNS sensory abnormalities, respectively. In case of disagreement, they reviewed the examinations together, until a shared decision was reached.

### Statistical analysis

In order to analyze difference of N20p-P25p amplitude related to age, gender and groups of patients with or without epilepsy/myoclonus, CNS and PNS sensory pathways abnormalities, Kruskall-Wallis and Mann– Whitney U tests were used. To analyze difference of N20p-P25p related to age, we divided the sample in age-related quartiles. Patients without ENG were excluded from the comparison of N20p-P25p amplitude; viceversa, for the cluster analysis, we assumed these patients were without PNS sensory abnormalities, for the normality of the peripheral response in the SEPs examination and the lacking of sensory symptoms. To identify the natural occurrence of subgroups of patients, a two-step cluster analysis with Schwarz’s Bayesian Criterion was applied considering age, presence of epilepsy/myoclonus, presence of abnormalities in CNS and/or PNS pathways and left and right SEP amplitudes. This is a hybrid approach to determine the number of clusters based on a statistical measure of fit and to use both categorical and continuous variables at the same time. To verify whether the clusters were different from one another, we performed statistical analysis using Kruskall-Wallis and chi-square tests with the cluster group as between-subjects factor. All of the statistical analyses were carried out using Statistical Package for the Social Sciences software, version 27 (SPSS Inc., Chicago, IL, U.S.A); p-values of < 0.05 were considered statistically significant.

## RESULTS

### Characteristics of the sample and descriptive statistics

All the data are summarized in table 2, figure 1 and figure 2.

**Table 2.** Data summary grouped by age-related quartiles and by etiological groups.

|  | Numb<br>er of<br>patient<br>s | Age<br>(years) | Sex<br>(M/F) | N20p-P25p<br>amplitude (μV) |  | Sensory<br>CNS<br>abnormaliti<br>es (Y/N) | Sensory PNS<br>abnormalities<br>(Y/N/undetermin<br>ed) | Presence<br>of<br>epilepsy<br>and/or<br>myoclon<br>us (Y/N) |
| --- | --- | --- | --- | --- | --- | --- | --- | --- |
|  |  |  |  | Left | Right |  |  |  |
| Age quartiles |  |  |  |  |  |  |  |  |
| First | 79 | 14-46 | 33/46 | 9.9±4.3 | 9.8±3.7 | 21/58 | 6/38/35 | 19/60 |
| Second | 72 | 47-59 | 29/43 | 8.6±2.5 | 9.0±2.6 | 23/49 | 10/33/29 | 8/64 |
| Third | 77 | 60-68 | 30/47 | 9.6±3.0 | 9.3±3.2 | 35/42 | 17/41/19 | 7/70 |
| Fourth | 80 | 69-91 | 31/49 | 9.4±3.0 | 9.9±3.6 | 42/38 | 19/37/24 | 6/74 |
|  | 308 | 14-91 | 123/185 | 9.4±3.3 | 9.5±3.3 | 122/186 | 52/151/105 | 40/268 |
| Etiological groups |  |  |  |  |  |  |  |  |
| CNS-IMM | 32 | 45.6±17.9 | 6/26 | 9.1±1.7 | 9.0±3.2 | 18/14 | 1/4/27 | 2/30 |
| CNS-LEUCO | 12 | 27.5±18.8 | 9/3 | 9.8±3.9 | 9.5±2.5 | 8/4 | 2/10/0 | 4/8 |
| CNS-TUM | 10 | 57.4±16.0 | 5/5 | 8.0±2.5 | 8.3±3.2 | 7/3 | 1/2/7 | 1/9 |
| CNS-VASC | 7 | 63.1±16.6 | 2/5 | 9.7±4.6 | 9.8±1.2 | 4/3 | 0/1/6 | 0/7 |
| COG.DIS | 23 | 66.9±9.0 | 10/13 | 9.1±1.6 | 9.2±2.3 | 6/17 | 1/8/14 | 10/13 |
| EPI-MYO | 20 | 28.4±19.4 | 8/12 | 13.9±6.7 | 12.1±5.6 | 4/16 | 3/6/11 | 20/0 |
| HSP | 10 | 43.7±21.6 | 6/4 | 10.7±4.1 | 8.9±3.3 | 1/9 | 0/7/3 | 0/10 |
| MND | 49 | 62.4±13.2 | 16/33 | 8.5±2.0 | 8.7±2.2 | 16/32 | 4/45/0 | 0/49 |
| MYOP | 13 | 56.2±19.9 | 6/7 | 8.5±1.4 | 9.1±1.9 | 3/10 | 1/12/0 | 0/13 |
| NEURO P | 18 | 62.3±20.2 | 11/7 | 9.1±2.2 | 9.5±2.5 | 5/13 | 18/0/0 | 0/18 |
| PARK | 28 | 69.4±8.1 | 11/17 | 9.8±4.3 | 10.1±3.3 | 11/17 | 4/15/9 | 2/26 |
| SCA | 8 | 60.0±5.9 | 3/5 | 9.3±1.6 | 7.9±3.0 | 7/1 | 1/4/3 | 2/6 |
| SPINE | 32 | 56.4±18.9 | 13/19 | 9.4±2.6 | 9.8±4.1 | 17/15 | 10/14/8 | 1/31 |
| OTHER-NEU | 22 | 53.0±18.8 | 9/13 | 9.2±2.5 | 9.3±3.0 | 9/13 | 2/11/9 | 0/22 |
| NON-ORGANIC | 9 | 54.3±16.7 | 1/8 | 9.0±3.7 | 11.6±4.8 | 0/9 | 0/4/5 | 0/9 |
| UND/UNK | 15 | 58.7±17.6 | 7/8 | 9.2±3.1 | 10.2±3.6 | 5/10 | 3/3/9 | 0/15 |
|  | **308** | **55.3±19.7** | **123/185** | **9.4±3.3** | **9.5±3.3** | **122/186** | **52/151/105** | **40/268** |

**Figure 1:**
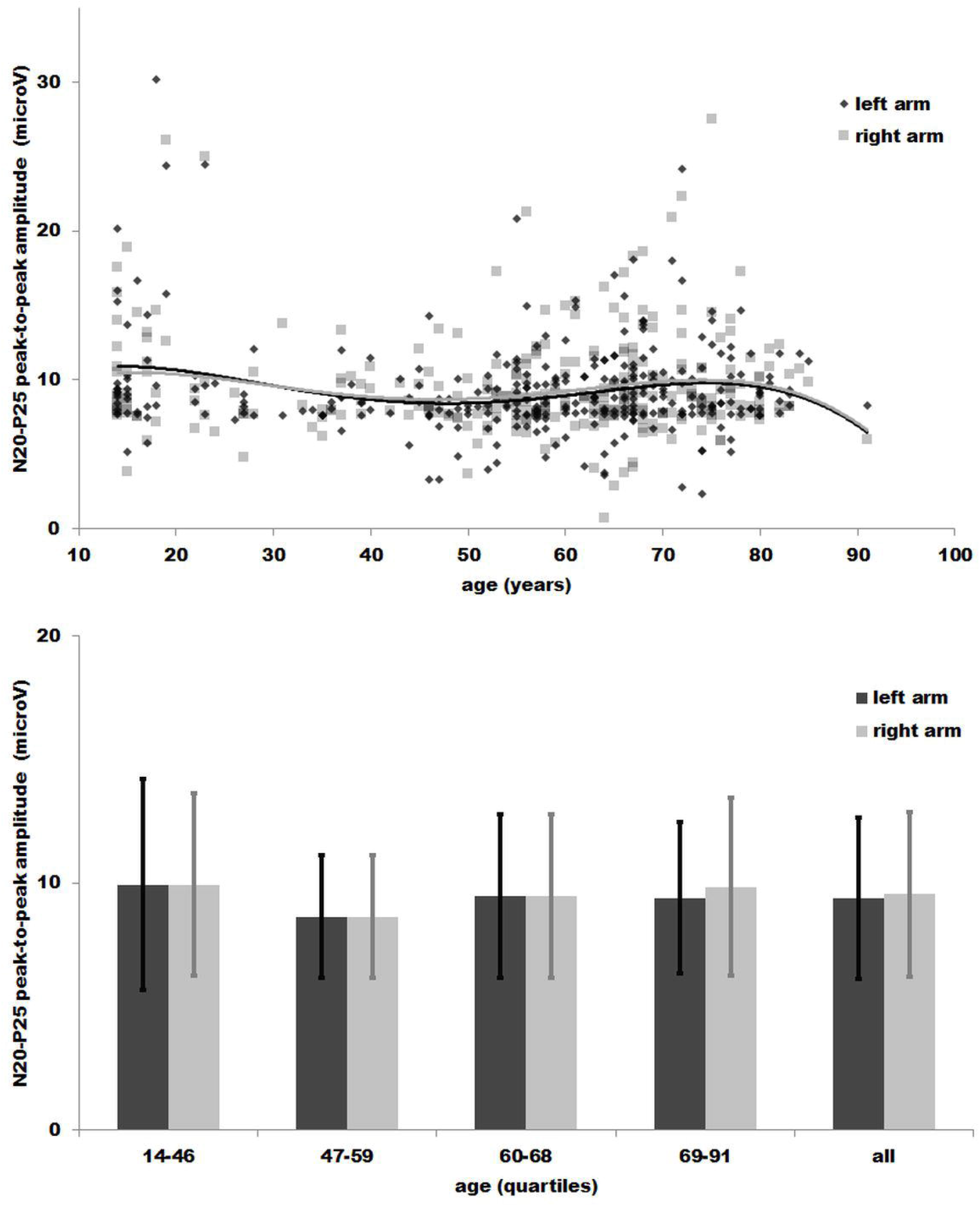
single data of the N20p-P25p amplitude for the left and right median nerve stimulation related to the age of patients are represented as dark grey diamonds and light grey squares, respectively; in the same colors, the means and the standard deviations of the N20p-P25p amplitude for the left and right median nerve are grouped in four quartiles of age.

**Figure 2:**
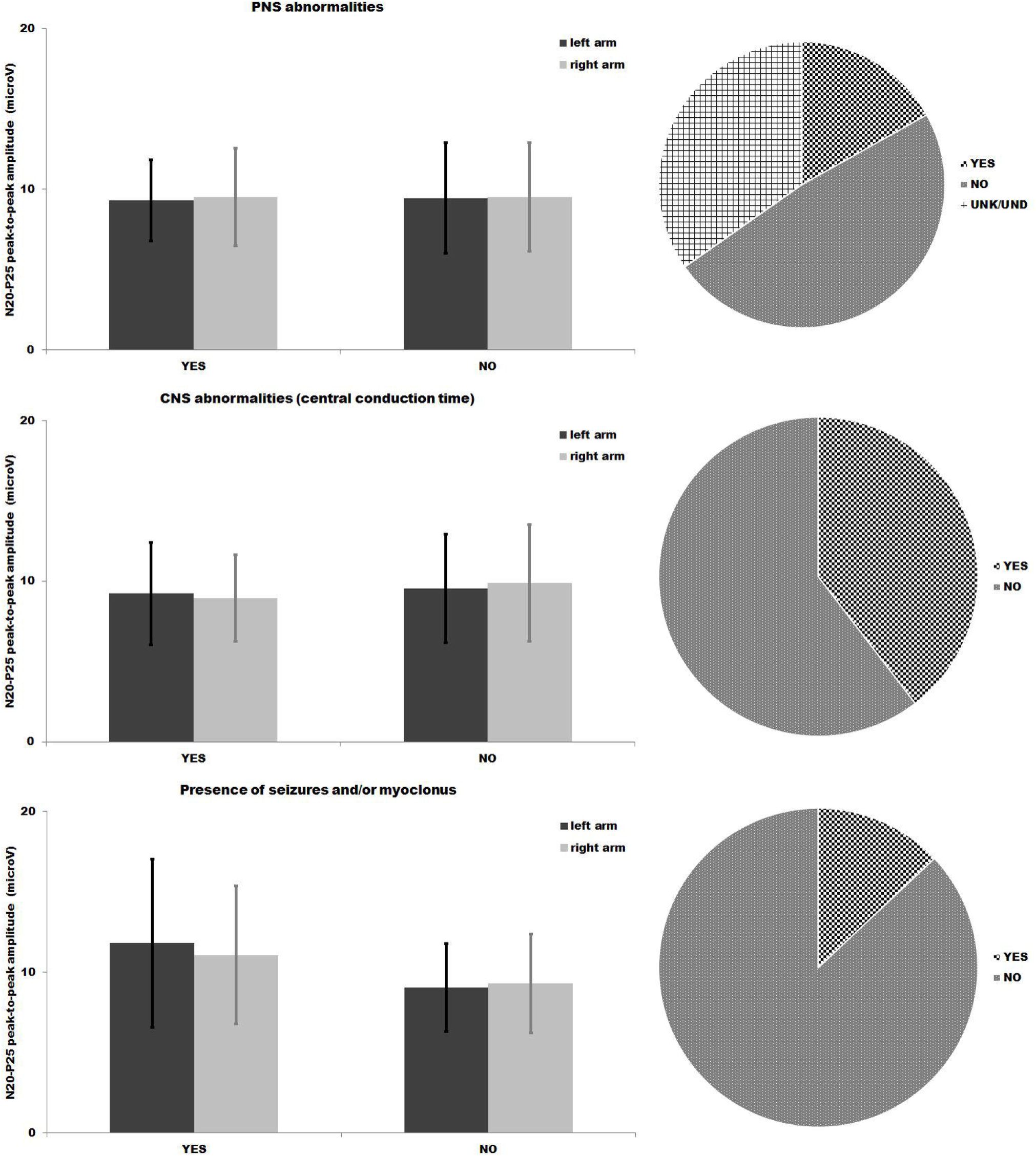
the percentage of patients with PNS and CNS abnormalities of the sensory system and of patients with epilepsy/myoclonus are represented together with the means and the standard deviations of the N20p-P25p amplitude for the left and right median nerve stimulations.

We identified 308 patients (123 males, mean age 55.2 years, range 14-91 years) with SEPIA. Mean N20p-P25p was 9.4 ± 3.3 and 9.5 ± 3.3 µV for left and right median nerve stimulation, respectively. A minority of them showed abnormalities of the CNS or PNS sensory pathways (39.6% and 16.6%, respectively). Epilepsy and/or myoclonus occurred only in the 13.0% of the patients.

The main group of patients were MND (15,9% of the total), followed by CNS-IMM and SPINE (for both, 10.3% of the total), PARK (9.1% of the total), COG.DIS (7.5% of the total), EPI-MYO (6.2% of the total) and NEUROP (5.8% of the total). With the exception of the OTHER-NEU group (7.1% of the total), a group composed of patients with various etiologies, no other group reached 5% of the total. Patients with unknown/undetermined diagnosis were 4.9% of the total.

### Statistical analysis

N20p-P25p were neither significantly different in presence of CNS alterations (left: U=6411, p=0.779; right: U=6492, p=0.676) nor PNS alterations (left: U=11083, p=0.763; right: U=10338, p=0.201), while it was significantly higher in patients with epilepsy and/or myoclonus (left: 9.06±2.73 µV vs. 11.81±5.24 µV, U=3125, p<0.001; right: 9.28±3.08 µV vs. 11.07±4.29 µV, U=<u>3</u>731.5, p=0.002). See table 2 for details. The asymmetric distribution of the sample across ages caused different interval of ages for the quartiles: 14-46 (N=79, 26.3±11.7 years), 47-59 (N=72, 54.2±3.4 years), 60-68 (N=77, 64.7±2.5 years, 69-91 (N=80, 75.6±4.6 years). No differences in N20p-P25p were found among quartiles, (left: H(3)=5.92, p=0.115; right: H(3)=3.06, p=0.383, table 2).

Excluding groups consisting of less than 10 subjects or with unknown etiology, a statistically significant difference in SEP amplitudes between the different etiological was found only for left side (H(9) = 18.89, p = 0.026). Post-hoc test revealed that patients with myoclonus or epilepsy had greater left N20p-P25p amplitudes with respect to all the other groups.

### Clusters analysis

Data of cluster analysis are summarized in table 3 and figure 3.

**Table 3.**
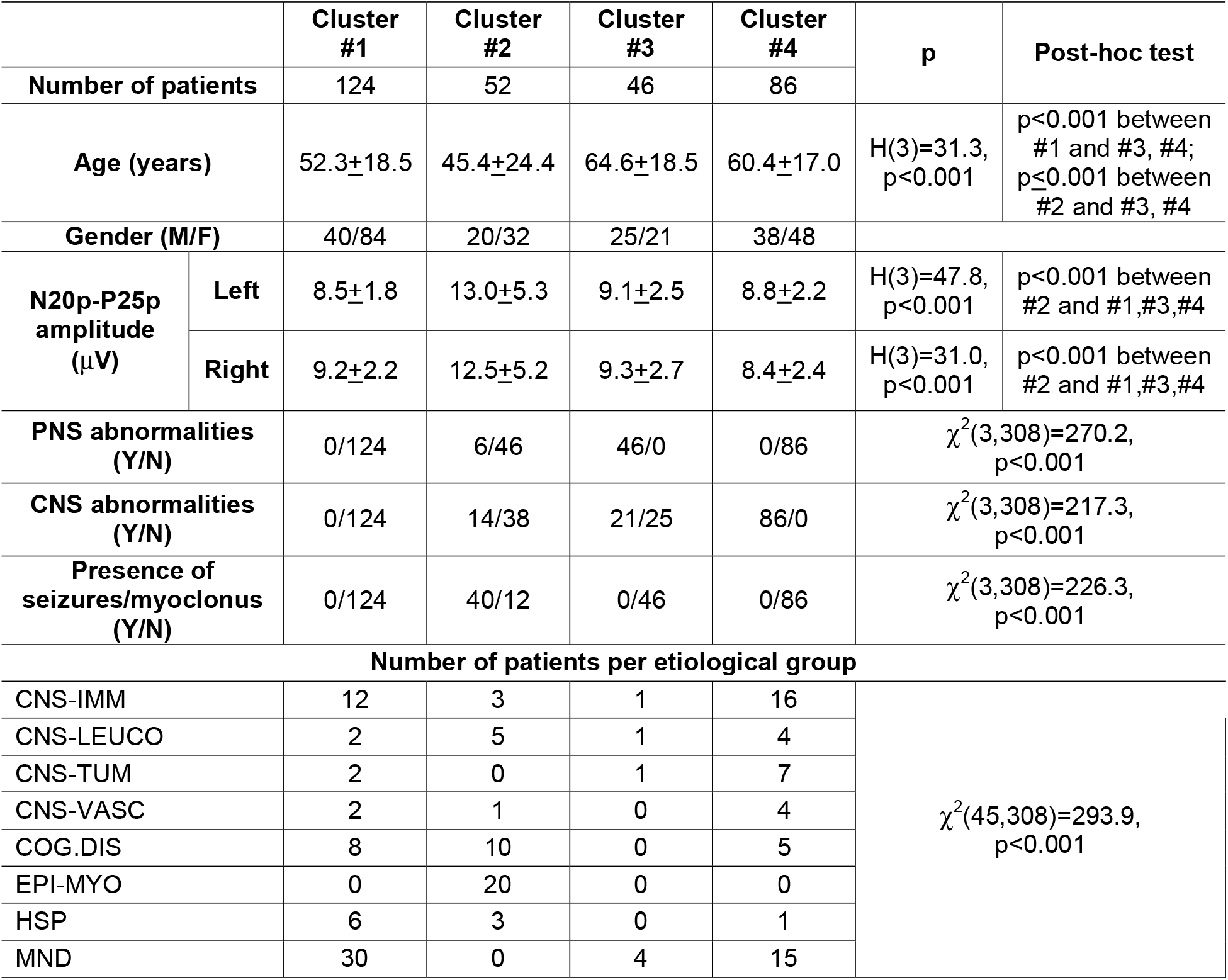
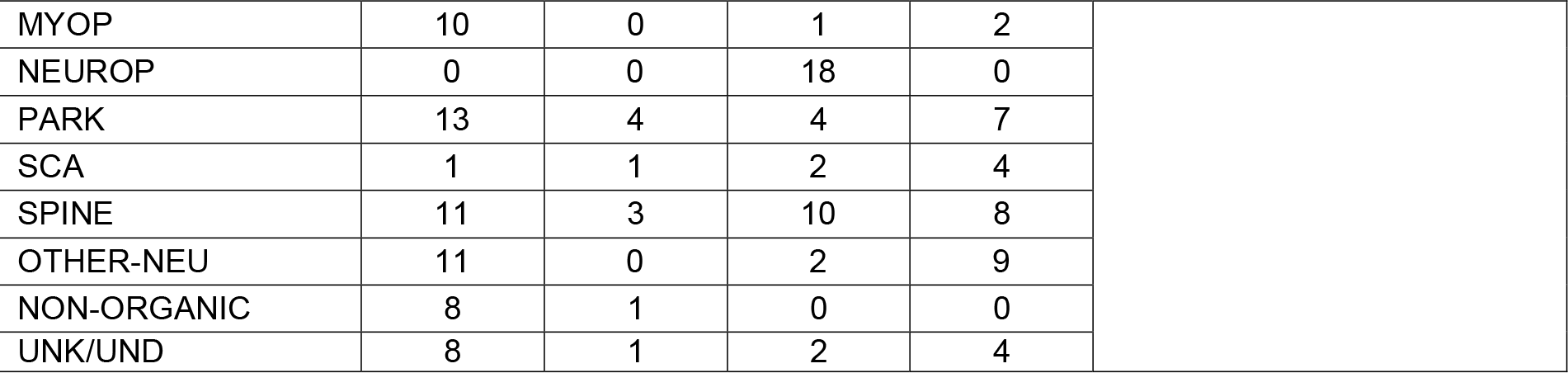
Cluster analysis.

|  |  | Cluster #1 | Cluster #2 | Cluster #3 | Cluster #4 | p | Post-hoc test |
| --- | --- | --- | --- | --- | --- | --- | --- |
| Number of patients |  | 124 | 52 | 46 | 86 |  |  |
| Age (years) | | 52.3±18.5 | 45.4±24.4 | 64.6±18.5 | 60.4±17.0 | H(3)=31.3, p<0.001 | p<0.001 between #1 and #3, #4; p≤0.001 between #2 and #3, #4 |
| Gender (M/F) |  | 40/84 | 20/32 | 25/21 | 38/48 |  |  |
| N20p-P25p amplitude (μV) | Left | 8.5±1.8 | 13.0±5.3 | 9.1±2.5 | 8.8±2.2 | H(3)=47.8, p<0.001 | p<0.001 between #2 and #1,#3,#4 |
| | Right | 9.2±2.2 | 12.5±5.2 | 9.3±2.7 | 8.4±2.4 | H(3)=31.0, p<0.001 | p<0.001 between #2 and #1,#3,#4 |
| PNS abnormalities (Y/N) | | 0/124 | 6/46 | 46/0 | 0/86 | $\chi^2$(3,308)=270.2, p<0.001 | |
| CNS abnormalities (Y/N) | | 0/124 | 14/38 | 21/25 | 86/0 | $\chi^2$(3,308)=217.3, p<0.001 | |
| Presence of seizures/myoclonus (Y/N) | | 0/124 | 40/12 | 0/46 | 0/86 | $\chi^2$(3,308)=226.3, p<0.001 | |
| Number of patients per etiological group |  |  |  |  |  |  |  |
| CNS-IMM | | 12 | 3 | 1 | 16 | $\chi^2$(45,308)=293.9, p<0.001 | |
| CNS-LEUCO |  | 2 | 5 | 1 | 4 |  |  |
| CNS-TUM |  | 2 | 0 | 1 | 7 |  |  |
| CNS-VASC |  | 2 | 1 | 0 | 4 |  |  |
| COG.DIS |  | 8 | 10 | 0 | 5 |  |  |
| EPI-MYO |  | 0 | 20 | 0 | 0 |  |  |
| HSP |  | 6 | 3 | 0 | 1 |  |  |
| MND |  | 30 | 0 | 4 | 15 |  |  |

|  |  |  |  |  |
| --- | --- | --- | --- | --- |
| MYOP | 10 | 0 | 1 | 2 |
| NEUROP | 0 | 0 | 18 | 0 |
| PARK | 13 | 4 | 4 | 7 |
| SCA | 1 | 1 | 2 | 4 |
| SPINE | 11 | 3 | 10 | 8 |
| OTHER-NEU | 11 | 0 | 2 | 9 |
| NON-ORGANIC | 8 | 1 | 0 | 0 |
| UNK/UND | 8 | 1 | 2 | 4 |

**Figure 3:**
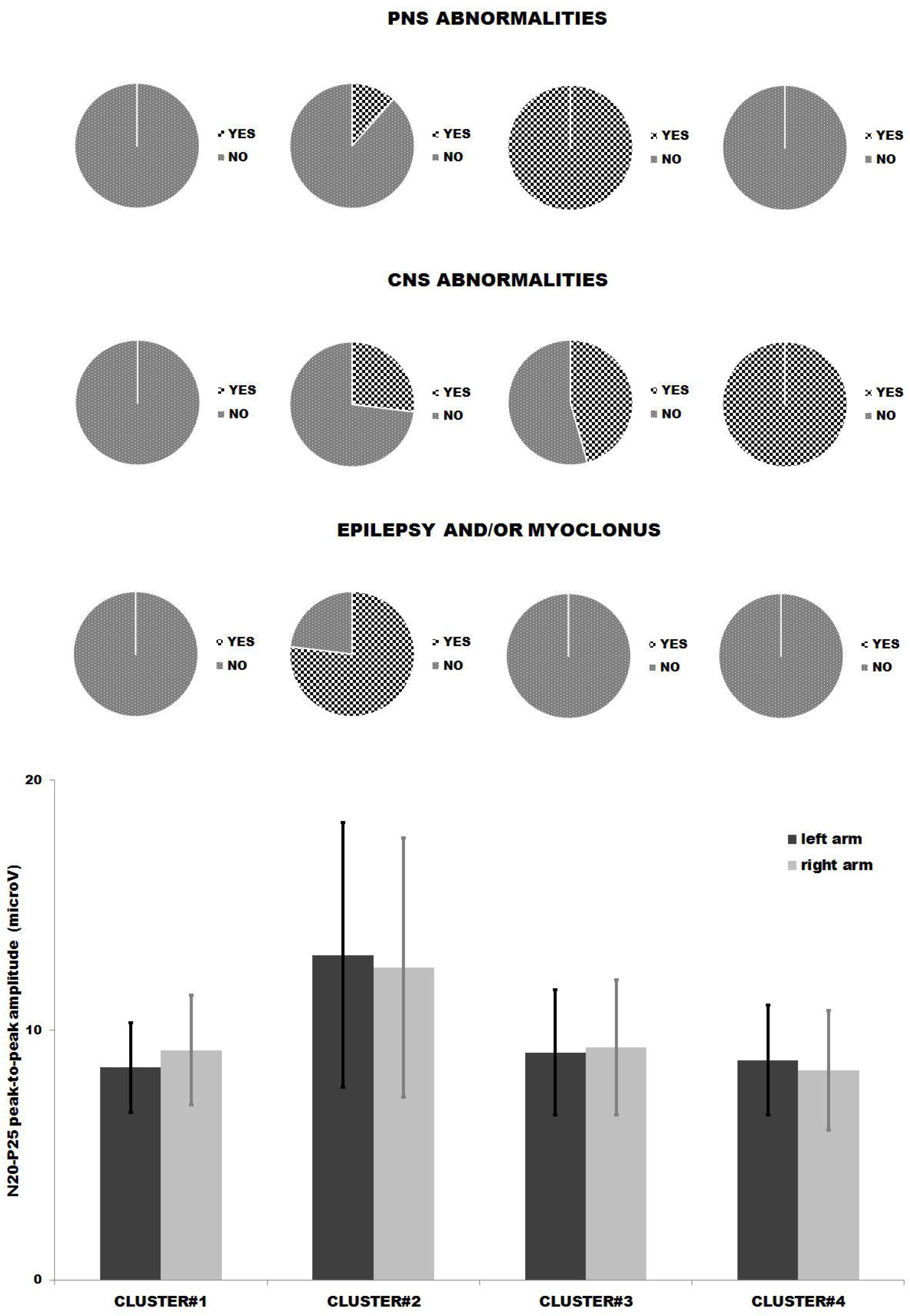
The different features of the four groups identified by the cluster analysis are shown: for each cluster, the percentage of patients with PNS and CNS abnormalities of the sensory system and of patients with epilepsy/myoclonus are represented in the upper part of the figure, together with the means and the standard deviations of the N20p-P25p amplitude for the left and right median nerve stimulations in the lower one (see also table 3). Cluster #2 showed significantly higher N20p-P25p values with respect to other groups (here represented by means of asterisks)

The two-steps cluster analysis identified four clusters, with the ratio largest/smallest clusters 2.70 (124: 46 patients). Patients included in the cluster#1 showed no abnormalities of CNS or PNS sensory pathways and absence of epilepsy and myoclonus. In cluster #2 most of the patients had epilepsy and/or myoclonus. All the patients included in cluster #3 showed PNS abnormalities of the sensory pathways; in cluster #4 all the patients showed abnormalities of CNS sensory pathways.

Post-hoc tests revealed that the differences were significant for the following parameters: both clusters #1 and #2 had patients younger than #3, #4, patients of cluster #2 had N20p-P25p higher than #1, #3 and #4, while there were no significant differences in the male/female ratio between the clusters. Moreover, clusters differed from each other for the presence/absence of CNS and PNS abnormalities, epilepsy and/or myoclonus and for the composition of patients.

## DISCUSSION

### Characteristics of the whole sample and statistical analysis

Characteristics of our sample were comparable to similar studies,[18,19] even if there was a higher prevalence of SEPIA in our group. The mean age of our population was similar to [19], while patients included in [18] were older. The predominance of females in our sample was probably related to the higher prevalence of some diseases, as autoimmune diseases, in the female population. Anyway, no significant differences of N20p-P25p related to age or sex and between patients with or without CNS or PNS sensory abnormalities were found.

In literature the term “SEPIA” and “giant SEPs” are indifferently used. Despite in former works reporting myoclonic patients, the term giant SEPs was restricted to a N20p-P25p reaching very high values (>15 µV), [12,13] in more recently ones [18,19] the same nomenclature has also been adopted for much smaller SEP amplitude. We think therefore that the term “giant” is more suitable for very enlarged SEPs amplitude, usually related to the presence of myoclonus, while “increased” can be referred to SEPs with only a mild increase with respect to the normative data; this subtle difference is not only lexical, because it might imply different pathophysiological conditions. In order to avoid misunderstandings, in our study we preferred to use SEPIA, because we included both myoclonic and non-myoclonic patients and because we could not to propose a limit value to differentiate between the “increased” and the “giant” SEPs (even considering confounding factors which we could not disentangle retrospectively, such as the intake of antiepileptic drugs). We found that myoclonic/epileptic patients had a significantly higher N20p-P25p with respect to other patients, confirming that an uncommonly large, “giant” cortical response to somatosensory input is usually related to the presence of myoclonus/epilepsy.

In the same line, according to our interpretation of the cluster analysis, SEPIA could be related to different pathophysiological conditions: a “strong” cortical hyperexcitability reflected in the very large N20p-P25p, in cortical myoclonus and, albeit to a lesser extent, epilepsy, as in patients of #2; “mild” cortical hyperexcitability related to a degenerative process involving cortex, as in patients of #1 or to plastic (maladaptive) cortical changes following PNS or CNS abnormalities of sensory pathways, as in patients of #3 or #4, respectively. Patients included in #2 were associated with cortical hyperexcitability for a long time, as for PMEs and other myoclonic[6-7,15] or epileptic syndromes,[12] prion diseases, [10] and some forms of parkinsonism with myoclonus.[20] In these patients, SEPIA are related to an abnormal response to the input occurring in the somatosensory and motor primary cortex due to their “intrinsic” hyperexcitability,[21] further enhanced by a cortical-subcortical (thalamic) loop.[22] This pathophysiological aberrant circuitry is also involved in the genesis of the reflex cortical myoclonus.[6-7,12,22]

In patients included in # 3 and # 4, SEPIA are related to plastic changes in somatosensory cortex following alterations in the body representation, due to peripheral and/or central body-brain pathways abnormalities. As demonstrated in studies based on functional neuroimaging, a remapping of the cortical somatotopy is present in patients following limb amputation[23] or brachial plexus avulsion,[24] in spinal cord injury,[25] in MS,[26] in stroke,[27] and in neuropathic painful syndromes.[28] The plastic changes to sensory deprivation, with deafferented areas replaced by neighbouring representations, result in a remodelling of the boundaries of the body map of the somatosensory cortex.[25] Some works even stated that these plastic changes could be maladaptive [25,29], thus contributing to secondary pathological conditions, such as hyperalgesia, allodynia, and neuropathic pain and phantom sensations.[23] To date, little is known about the link between SEPIA, (maladaptive) plastic changes after damage of the sensory pathways and the occurrence of sensory symptoms. Interestingly, temporary deafferentiation determines increase of earlier components of SEPs, included N20 and P25.[30-31] Hence, we could speculate that partial deafferentation of somatosensory cortex may produce a similar increase in the patients of clusters #3 and #4. Indeed, #3 and #4 were populated by patients with peripheral neuropathies (NEUROP) and with abnormalities of the ascending sensory pathways (SPINE, SCA, CNS-LEUCO, most CNS-IMM groups), respectively.

In cluster #1 no patients showed abnormalities of sensory pathways, myoclonus or epilepsy. Hence, we assume that in cluster #1 SEPIA were related to pathophysiological (degenerative?) changes occurring at the cortical or cortico-subcortical level, as for the patients included in the MND and PARK groups (included MSA, PSP) and with primary dementia. Few studies assessed the presence of SEPIA in neurodegenerative diseases: patients affected by ALS had a larger N20p-P25p, with respect to the healthy subjects; moreover, median survival time after examination was shorter in the group of ALS with N20p-P25p greater than 8 µV.[17] This large N20p-P25p was interpreted as a sign of cortical hyperexcitability in the sensorimotor cortical areas in the first stages of ALS, due to the early loss of cortical interneurons, preceding the loss of the upper motor neuron. SEPIA was found in PSP[15] and in MSA;[14] again, this finding probably reflected a mild form of cortical hyperexcitability. To our knowledge, no other studies focused on SEPIA, even if it is well known that unbalancing of the cortical excitability is present in several neurodegenerative disorders, including dementia Alzheimer type, sporadic ALS, fronto-temporal dementia, PD and parkinsonisms,[32] in MS [33] and in genetic form of ALS/FTD patients.[34] All together these data corroborate our hypothesis. At individual level, two or more processes could contribute in determining presence of SEPIA, especially for diseases in which both a degenerative cortical process and an involvement of CNS sensory pathways are expected, such as ALS, MSA, MS.

Not all patients fit fully into our interpretative model; some patients in #2 showed neither myoclonus nor epilepsy: the inclusion in this cluster probably depended by the large N20p-P25p amplitude which have influenced the composition of #2. Then, sensory abnormalities in myopathies and in neuromuscular junction diseases are largely underestimated, even in neurophysiological examinations.[35] Hence, the patients in group MYOP probably showed increased N20p-P25p as a result of plastic changes related to sensory abnormalities, as example proprioceptive, due to the myopathy; their inclusion in #1 was due to the fact that these abnormalities were unrevealed by conventional neurophysiological examination.

### The possible use of SEPs increased in amplitude in the clinical context

In our case series, different neurological disorders were represented, validating the hypothesis that SEPIA could not be considered *per se* as neither a marker of a specific disease, nor of a sign (myoclonus) and nor to a definite pathophysiological process. Nevertheless, some useful information can be obtained for clinical use by the presence of SEPIA: our data open a window in considering SEPIA more than a “simple” neurophysiological sign of cortical myoclonus and, even considering that SEP examination is a low-cost, largely used technique, we propose it as a first-level diagnostic tool for all the conditions in which a degenerative or plastic rearrangement of somatosensory cortex can be hypothesized; especially in non-myoclonic patients, the occurrence of a large N20p-P25p should induce clinicians to further investigate a possible degenerative process involving somato(motor) cortex considering the clinical, neurophysiological and neuroradiological picture as a whole.In ALS patients, SEPIA highlight subclinical early cortical involvement and a worse prognosis;[17] in parkinsonisms, SEPIA are probably more frequent in MSA and PSP than in PD; in cognitive disorders, very large N20p-P25p strongly relates to a prion disease (first of all CJD).

Furthermore, the existence of patients with functional disorders with SEPIA, was certainly the most counterintuitive finding of our study, because it is problematic to imagine for them adaptive or degenerative cortical changes. Anyway, other Authors[19] reported in their sample some cases which could be referred to “functional” disorders. Gurses et al. [36] reported a single case-report of a very large increased of the P25 component in a patient with a motor conversion disorder, while another child showed transient disappearance of cortical response to the stimulation of the lower limbs associated to transient paraparesis;[37] moreover, imagined paralysis decreased SEP amplitude even in healthy subjects.[38] Although contradictory, all these studies endorse the hypothesis that changes in bodily self-representation can modulate even “low-level” somatosensory processing.[39] Further studies need to clarify if SEPIA could be a hallmark of modified self-awareness and/or self-representation in functional patients. Finally, it has to be considered if SEPIA may become an index of plasticity in treatments aimed to restore cortical functioning.

### Limitations

Some limitations must be accounted; the prevalence of SEPIA found in our study may not reflect the real one in the neurological disorders: we more frequently included diseases for which the execution of SEPs was integrated in the routinely diagnostic work-up. This sample bias depended on the retrospective nature of our study.

Again, the division of patients into various “diagnostic groups” could be disputed; we did so to facilitate the interpretation of results, even though we had accounted for diagnostic groups in neither statistical nor cluster analysis. Therefore, although questionable, this subdivision of patients did not influence the methods and results.

Finally, we took into consideration only the earliest cortical components of SEPs, while previous works stressed also later components and the dynamics of the recovery of SEPs could be abnormal [7,15]. However, our primary aim was to investigate about the significance of a finding (i.e., increased N20p-P25p) usually underestimated in the daily diagnostic work-up.

## Data Availability

All data produced in the present study are available upon reasonable request to the authors

## REFERENCES

1. Cruccu G, Aminoff MJ, Curio G, et al. Recommendations for the clinical use of somatosensory-evoked potentials. Clin Neurophysiol. 2008; 119:1705–1719.

2. Guérit JM, Amantini A, Amodio P, et al. Consensus on the use of neurophysiological tests in the intensive care unit (ICU): electroencephalogram (EEG), evoked potentials (EP), and electroneuromyography (ENMG). Neurophysiol Clin. 2009;39:71–83.

3. Koutlidis RM, Ayrignac X, Pradat PF, et al. Segmental somatosensory-evoked potentials as a diagnostic tool in chronic inflammatory demyelinating polyneuropathies, and other sensory neuropathies. Neurophysiol Clin. 2014;44:267–80.

4. Dawson GD. Cerebral responses to electrical stimulation of peripheral nerve in man. J Neurol Neurosurg Psychiatry 1947;10:134–140.

5. Rothwell JC, Obeso JA, Marsden CD. On the significance of giant somatosensory evoked potentials in cortical myoclonus. J Neurol Neurosurg Psychiatry. 1984;47:33–42.

6. Shibasaki H, Yamashita Y, Neshige R, et al. Pathogenesis of giant somatosensory evoked potentials in progressive myoclonic epilepsy. Brain. 1985;108:225–40.

7. Cantello R, Gianelli M, Civardi C, et al. Focal subcortical reflex myoclonus. A clinical and neurophysiological study. Arch Neurol. 1997;54(2):187–96.

8. Takeuchi H, Touge T, Miki H, et al. Electrophysiological and pharmacological studies of somatosensory reflex myoclonus. Electromyogr Clin Neurophysiol. 1992;32:143–54.

9. Plasmati R, Michelucci R, Forti A, et al. The neurophysiological features of benign partial epilepsy with rolandic spikes. Epilepsy Res Suppl. 1992;6:45–8.

10. Matsunaga K, Uozumi T, Akamatsu N, et al. Negative myoclonus in Creutzfeldt-Jakob disease. Clin Neurophysiol. 2000;111:471–6.

11. Schmitt B, Thun-Hohenstein L, Molinari L, et al. Somatosensory evoked potentials with high cortical amplitudes: clinical data in 31 children. Neuropediatrics. 1994;25:78–84.

12. Canafoglia L, Ciano C, Panzica F, et al. Sensorimotor cortex excitability in Unverricht-Lundborg disease and Lafora body disease. Neurology. 2004;63(12):2309–15.

13. Visani E, Canafoglia L, Rossi Sebastiano D, et al. Giant SEPs and SEP-recovery function in Unverricht-Lundborg disease. Clin Neurophysiol. 2013;124:1013–8.

14. Rodriguez ME, Artieda J, Zubieta JL, et al. Reflex myoclonus in olivopontocerebellar atrophy. J Neurol Neurosurg Psychiatry. 1994;57:316–9.

15. Kofler M, Müller J, Reggiani L, et al. Somatosensory evoked potentials in progressive supranuclear palsy. J Neurol Sci. 2000;179:85–91.

16. Nanetti L, Sarto E, Castaldo A, et al. ANO10 mutational screening in recessive ataxia: genetic findings and refinement of the clinical phenotype. J Neurol. 2019;266:378–385.

17. Shimizu T, Bokuda K, Kimura H, et al. Sensory cortex hyperexcitability predicts short survival in amyotrophic lateral sclerosis. Neurology. 2018;90:e1578–e1587.

18. Martín-Palomeque G, Castro-Ortiz A, Pamplona-Valenzuela P, et al. Large Amplitude Cortical Evoked Potentials in Nonepileptic Patients. Reviving an Old Neurophysiologic Tool to Help Detect CNS Pathology. J Clin Neurophysiol. 2017;34:84–91.

19. Horlings CGC, Kofler M, Hotter A, et al. The clinical meaning of giant somatosensory evoked potentials of the median nerve. Clin Neurophysiol. 2020;131:1495–1496.

20. Defebvre L. Myoclonus and extrapyramidal diseases. Neurophysiol Clin. 2006;36:319–25.

21. Hitomi T, Ikeda A, Matsumoto R, et al. Generators and temporal succession of giant somatosensory evoked potentials in cortical reflex myoclonus: epicortical recording from sensorimotor cortex. Clin Neurophysiol. 2006;117:1481–6.

22. Manganotti P, Tamburin S, Zanette G, et al. Hyperexcitable cortical responses in progressive myoclonic epilepsy: a TMS study. Neurology. 2001;57:1793–9.

23. Makin TR, Flor H. Brain (re)organisation following amputation: Implications for phantom limb pain. Neuroimage. 2020;218:116943.

24. Ramachandran VS, Hirstein W. The perception of phantom limbs. The D. O. Hebb lecture. Brain. 1998;121:1603–30.

25. Leemhuis E, De Gennaro L, Pazzaglia AM. Disconnected Body Representation: Neuroplasticity Following Spinal Cord Injury. J Clin Med. 2019;8:2144.

26. Filippi M, Preziosa P, Rocca MA. Brain mapping in multiple sclerosis: Lessons learned about the human brain. Neuroimage. 2019;190:32–45.

27. Winship IR, Murphy TH. Remapping the somatosensory cortex after stroke: insight from imaging the synapse to network. Neuroscientist. 2009;15:507–24.

28. Gustin SM, Peck CC, Cheney LB, et al. Pain and plasticity: is chronic pain always associated with somatosensory cortex activity and reorganization? J Neurosci. 2012;32:14874–84.

29. Oouchida Y, Sudo T, Inamura T, et al. Maladaptive change of body representation in the brain after damage to central or peripheral nervous system. Neurosci Res. 2016;104:38–43.

30. Tinazzi M, Rosso T, Zanette G, et al. Rapid modulation of cortical proprioceptive activity induced by transient cutaneous deafferentation: neurophysiological evidence of short-term plasticity across different somatosensory modalities in humans. Eur J Neurosci. 2003;18:3053–60.

31. Dubois JD, Poitras I, Voisin JIA, et al. Effect of pain on deafferentation-induced modulation of somatosensory evoked potentials. PLoS One. 2018;13:e0206141.

32. Vucic S, Kiernan MC. Transcranial Magnetic Stimulation for the Assessment of Neurodegenerative Disease. Neurotherapeutics. 2017;14:91–106.

33. Chaves AR, Wallack EM, Kelly LP, et al. Asymmetry of Brain Excitability: A New Biomarker that Predicts Objective and Subjective Symptoms in Multiple Sclerosis. Behav Brain Res. 2019; 359:281–291.

34. Starr A, Sattler R. Synaptic dysfunction and altered excitability in C9ORF72 ALS/FTD. Brain Res. 2018;1693:98–108.

35. Leon-Sarmiento FE, Leon-Ariza JS, Prada D, et al. Sensory aspects in myasthenia gravis: A translational approach. J Neurol Sci. 2016;368:379–88.

36. Gürses N, Temuçin CM, Lay Ergün E, et al. Konversiyon bozukluğunda uyarilmiş potansiyeller ve beyin kan akimi değişiklikleri: olgu sunumu ve gözden geçirme [Evoked potentials and regional cerebral blood flow changes in conversion disorder: a case report and review]. Turk Psikiyatri Derg. 2008;19:101–7. Turkish.

37. Yu T, Ye J, Deng Y, et al. Abnormal somatosensory evoked potentials in a child with motor conversion disorder: A case report. Psychiatry Clin Neurosci. 2021;75:319–320.

38. Palluel E, Falconer CJ, Lopez C, et al. Imagined paralysis alters somatosensory evoked-potentials. Cogn Neurosci. 2020;11:205–215.

39. Aspell JE, Palluel E, Blanke O. Early and late activity in somatosensory cortex reflects changes in bodily self-consciousness: an evoked potential study. Neuroscience. 2012;216:110–22.

